# Vaccine uptake and SARS-CoV-2 antibody prevalence among 207,337 adults during May 2021 in England: REACT-2 study

**DOI:** 10.1101/2021.07.14.21260497

**Authors:** Helen Ward, Matt Whitaker, Sonja N Tang, Christina Atchison, Ara Darzi, Christl A Donnelly, Peter J. Diggle, Deborah Ashby, Steven Riley, Wendy S Barclay, Paul Elliott, Graham Cooke

## Abstract

**Background:** The programme to vaccinate adults in England has been rapidly implemented since it began in December 2020. The community prevalence of SARS-CoV-2 anti-spike protein antibodies provides an estimate of total cumulative response to natural infection and vaccination. We describe the distribution of SARS-CoV-2 IgG antibodies in adults in England in May 2021 at a time when approximately 7 in 10 adults had received at least one dose of vaccine.

**Methods:** Sixth round of REACT-2 (REal-time Assessment of Community Transmission-2), a cross-sectional random community survey of adults in England, from 12 to 25 May 2021; 207,337 participants completed questionnaires and self-administered a lateral flow immunoassay test producing a positive or negative result.

**Results:** Vaccine coverage with one or more doses, weighted to the adult population in England, was 72.9% (95% confidence interval 72.7-73.0), varying by age from 25.1% (24.5-25.6) of those aged 18 to 24 years, to 99.2% (99.1-99.3) of those 75 years and older. In adjusted models, odds of vaccination were lower in men (odds ratio [OR] 0.89 [0.85-0.94]) than women, and in people of Black (0.41 [0.34-0.49]) compared to white ethnicity. There was higher vaccine coverage in the least deprived and highest income households. People who reported a history of COVID-19 were less likely to be vaccinated (OR 0.61 [0.55-0.67]). There was high coverage among health workers (OR 9.84 [8.79-11.02] and care workers (OR 4.17 [3.20-5.43]) compared to non-key workers, but lower in hospitality and retail workers (OR 0.73 [0.64-0.82] and 0.77 [0.70-0.85] respectively) after adjusting for age and key covariates.

The prevalence of antibodies (weighted to the adult population of England and adjusted for test characteristics) was 61.1% (95% CI 60.9-61.4), up from 6.6% (5.4-5.7) in round 4 (27 October to 10 November 2020) and 13.9% (13.7-14.1) in round 5 (26 January to 8 February 2021). Prevalence (adjusted and weighted) increased with age, from 35.8% (35.1-36.5) in those aged 18 to 24 years, to 95.3% (94.6-95.9) in people 75 and over. Antibodies were 30% less likely to be detected in men than women (adjusted OR 0.69, 0.68-0.70), and were higher in people of Asian (OR 1.67 [1.58-1.77]), Black (1.55 [1.41-1.69]), mixed 1.17 [1.06-1.29] and other (1.37 [1.23-1.51]) ethnicities compared with white ethnicity. Workers in hospitality (OR 0.69 [0.63-0.74]) and retail (0.71 [0.67-0.75]) were less likely to have antibodies.

Following two doses of Pfizer-BioNTech vaccine, antibody positivity (adjusted for test performance) was 100% (100-100) at all ages except 80 years and older when it was 97.8% (95.9-99.6). For AstraZeneca positivity was over 90% up to age 69, and then 89.2% (88.5-89.9) in 70-79 year olds and 83.6% (78.5-88.3) in those aged 80 and over. Following a single dose of Pfizer-BioNTech positivity ranged from 100.0% (91.1-100.0) in those aged 18-29 to 32.2% (18.2-51.1) in those aged 70-79 years. For AstraZeneca this was 72.2% (68.5-75.9) in the youngest and 46.2% (40.0-52.7) in the oldest age group.

**Discussion:** The successful roll out of the vaccination programme in England has led to a high proportion of individuals having detectable antibodies, particularly in older age groups and those who have had two doses of vaccine. This is likely to be associated with high levels of protection from severe disease, and possibly from infection. Nonetheless, there remain some key groups with a lower prevalence of antibody, notably unvaccinated younger people, certain minority ethnic groups, those living in deprived areas and workers in some public facing employment. Obtaining improved rates of vaccination in these groups is essential to achieving high levels of protection against the virus through population immunity.

**Funding:** Department of Health and Social Care in England.

## Introduction

England has experienced a high and uneven burden of COVID-19 across the population, with high mortality and morbidity in care homes, in minority ethnic groups, and in deprived communities.(1). Prior to the launch of the COVID-19 vaccination programme the prevalence of IgG antibody to the SARS-CoV-2 spike protein provided an estimate of the cumulative burden of infection across the population.(2) The COVID-19 vaccination programme started in England in December 2020 with a phased roll-out based on recommendations from the UK Joint Committee on Vaccination and Immunization (JCVI), which built on the WHO Roadmap.(3,4). Once vaccination was introduced, antibody prevalence captured both prior infection and occurrence of antibodies following one or two vaccine doses.

Implementation of the vaccination programme in England has been rapid, with initial deployment of first doses speeded up by delaying the second dose to 12 weeks; by 15 May 2021 69% of adults in England had received their first and 39% their second dose of vaccine, and by 1 July 2021, these proportions were 85% and 63% respectively.(4) There is growing evidence that the programme, initially targeting older adults, health and care workers, care home residents and clinically extremely vulnerable people, is reducing transmission, hospitalisations and mortality.(5)

The REal-time Assessment of Community Transmission-2 (REACT-2) study is a community survey to measure the prevalence of antibodies to the SARS-CoV-2 virus among random samples of adults in England. We previously reported the extent and variation in antibody prevalence after the first wave of COVID-19 in England including the unequal risk by region, occupation and ethnicity.(2) Subsequent rounds of REACT-2 showed a decline in prevalence over the summer of 2020 as antibodies waned in the population,(6) followed by an increase following the second wave and the first few weeks of the vaccination programme.(7) Here we report the results of REACT-2 round 6 carried out between 12 and 25 May 2021 to measure antibody prevalence across the adult population in England.

## Methods

REACT-2 methods have been published elsewhere(2,8). Briefly, in each round of the study we invited a non-overlapping random community sample of adults aged 18 years or over in England, based on the National Health Service general practitioner registrations list. The sample was designed to provide approximately equal numbers of people in each of the 315 lower tier local authority (LTLA) areas in England. Those who registered were sent a Fortress lateral flow immunoassay (LFIA) test kit for SARS-CoV-2 antibody self-testing and asked to perform the test at home and complete a questionnaire, including reporting of their test result and uploading a photo of the completed test. Participants were also asked to report on COVID-19 vaccination, with dates. Survey instruments are available on the study website^1^. Those who had not received a vaccine dose were asked whether they had been invited to take part in the vaccination programme and their response. For those who had not yet been offered a vaccine we asked about their intention to accept. People who reported being unsure or who would decline vaccination were asked to select from a list of possible reasons for hesitancy(9) with the option also of providing free-text responses.

Round 6 adopted the design of previous rounds with the addition of a boosted sample of older adults to increase the power to detect whether the risk of becoming a case, being hospitalised or dying from COVID-19 differed between those testing positive and those testing negative on the LFIA following vaccination. Round 6 aimed for a total sample size of 240,000, with inclusion of 70,000 additional people in age groups 55-64 and 65-67 years. We estimated that this boosted sample of older age groups would give high probability (p=0.013) of detecting clinically important differences in outcomes (relative risk 0.5 for hospitalisation).

The LFIA used in REACT-2 detects immune responses to the S1 subunit protein targeted by available vaccines. It was selected following evaluation of performance characteristics (sensitivity and specificity) against pre-defined criteria for detection of IgG,(10) with extensive public involvement and user testing.(11) We estimated clinical sensitivity of the LFIA on finger-prick blood (self-read) for IgG antibodies following natural infection at 84.4% (70.5, 93.5) among RT-PCR confirmed cases in healthcare workers, and specificity 98.6% (97.1, 99.4) in pre-pandemic sera.(10,12)

Here we report data on vaccine coverage and results of antibody testing based on complete, validated data from round 6 received on 28 May 2021.

### Data analyses

Prevalence was calculated as the proportion of individuals with a positive IgG result on the LFIA. For analyses at population level (but not for individual vaccine response) we adjusted for test performance.(13) Prevalence estimates for antibodies and vaccine coverage at national level were weighted for age, sex, region, ethnicity and deprivation to account for the geographic sample design and for variation in response rates, so as to be representative of the population (18+ years) of England. Index of Multiple Deprivation 2019 (IMD) was used as a measure of relative deprivation, based on seven domains at local-area level across England (income, employment, education, health, crime, barriers to housing and services, and living environment).(14)

We used multivariable logistic regression to investigate the association of covariates with vaccine coverage and to estimate odds of antibody positivity adjusting for potential confounders. These included age, sex, days since vaccine and previous COVID-19 infection status, as well as interaction effects between age and sex.

Data were analysed using the statistical package R version 4.0.0.(15). We obtained research ethics approval from the South Central-Berkshire B Research Ethics Committee (IRAS ID: 283787), and Medicines and Healthcare products Regulatory Agency approval for use of the LFIA for research purposes only.

### Public involvement

The REACT Public Advisory Panel has provided regular review and revision of the study processes and results.

### Funding

This study was funded by the Department of Health and Social Care in England. The content of this manuscript and decision to submit for publication were the responsibility of the authors and the funders had no role in these decisions.

## Results

### Sample

From 12 to 25 May 2021, 207,337 adults reported an IgG positive or negative result from their self-test. Overall, 28% of all those invited and 81% of those who registered submitted a test result; 97.1% of those who attempted the test managed to complete it and report a positive or negative result. Further details on response patterns are provided in Supplementary Data (Supplementary Table S1).

### Vaccine coverage and confidence

There were high rates of vaccine coverage (weighted to be representative of the adult population in England) with 72.9% (95% confidence interval 72.7-73.0) reporting having had at least one dose by the time of the survey, varying by age from 25.1% (24.5-25.6) of those aged 18 to 24 years, to 99.2% (99.1-99.3) of those aged 75 years and older (Supplementary Table S2).

Figure 1 shows variation in odds of vaccination; coverage increased steeply with age in line with the vaccine roll-out, and odds adjusted for age were lower in men (odds ratio [OR] 0.89 [0.85-0.94]) than women. After adjustment, vaccine coverage was lower in people of Black (0.41 [0.34-0.49]), and higher in people of Asian (1.13 [1.01-1.27]) ethnicity, than white ethnicity. There was a gradient of coverage in relation to deprivation and household income, with higher coverage in the least deprived and highest income households (Supplementary Table S3). There was variation by region, with odds being lower in London (OR 0.86 [0.79-0.94]) and higher in the North West (OR 1.23 [1.13-1.35]) than the South East. At LTLA level, the weighted vaccine coverage varied between 43.4% and 87.5% (Figure 2, Supplementary Data File). People who reported a history of COVID-19 were less likely to be vaccinated (OR 0.61 [0.55-0.67]). There was high coverage among health and care workers with odds of 9.84 [8.79-11.02] and 4.17 [3.20-5.43] respectively compared to non-key workers, and odds were higher in education workers than other workers at 1.63 [1.50-1.76]. However, coverage was low in hospitality and retail workers compared to other workers (OR 0.73 [0.64-0.82] and 0.77 [0.70-0.85] respectively) (Supplementary Table S3).

**Figure 1.**
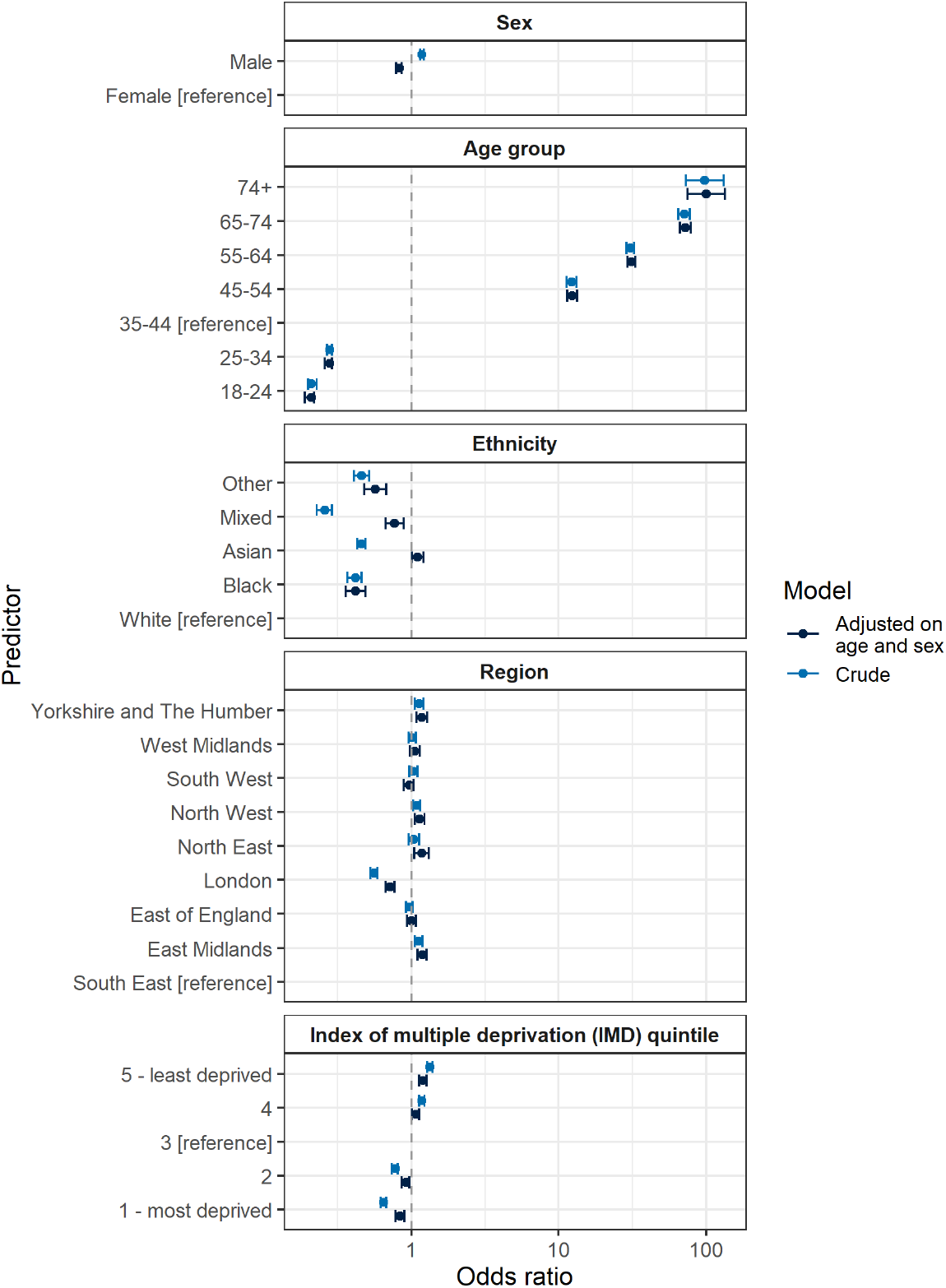
Logistic regression for vaccine coverage (one or more doses received, y/n) by key covariates. Legend: The plot shows odds ratios for logistic regression models with vaccination status as the binary outcome variable (one or more doses received, y/n) and a range of covariates as explanatory variables, crude and adjusted for age and sex. Data for this chart are in Supplementary Table S3

**Figure 2.**
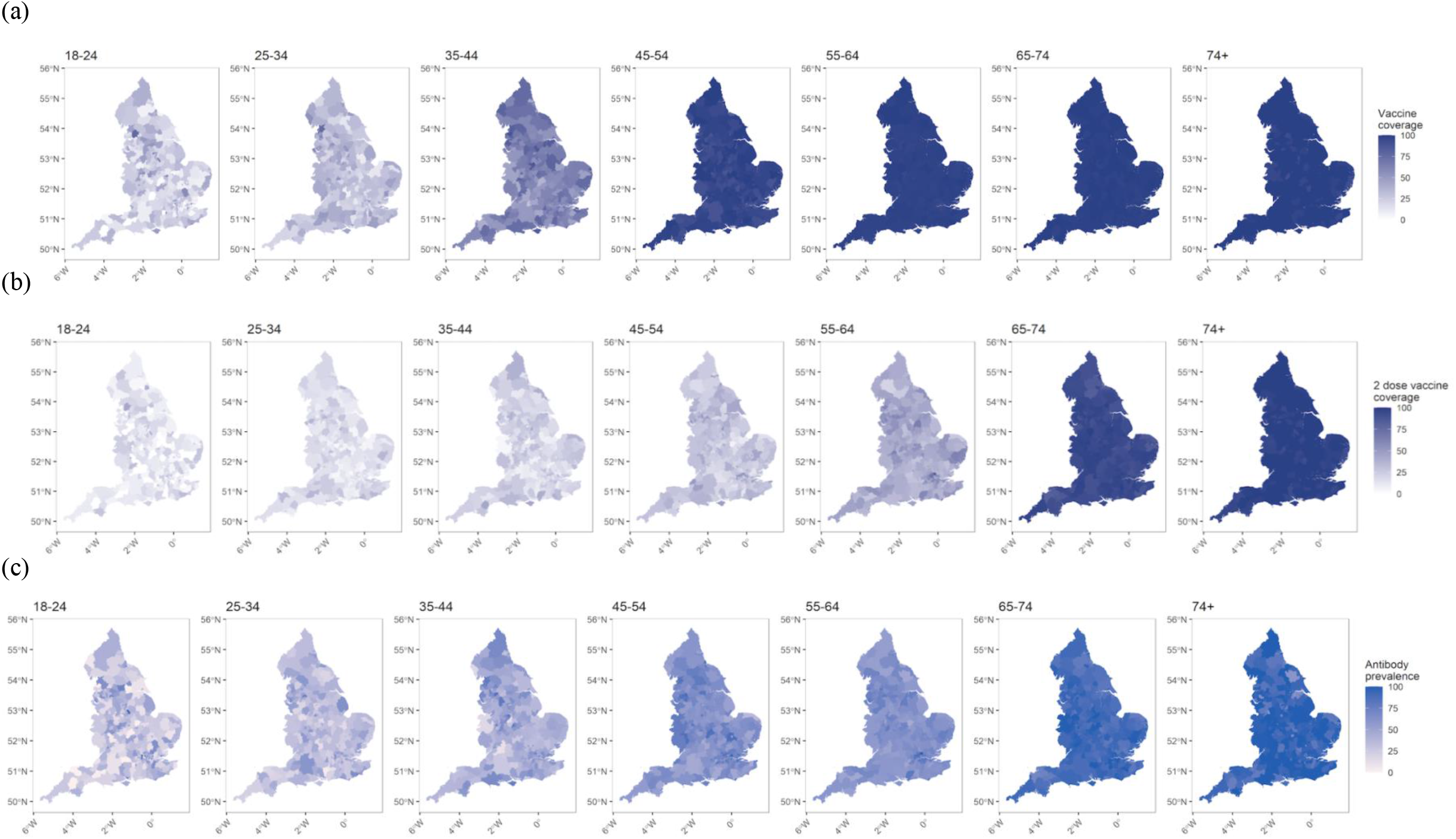
Maps of England showing variation at lower-tier local authority (LTLA) level by age for (a) antibody prevalence (adjusted and weighted), (b) proportion (weighted) with at least one vaccine dose, (c) proportion (weighted) double vaccinated.

Overall vaccine confidence was high, with 97.6%, [97.5-97.6] reporting having been vaccinated or intending to accept a vaccine when offered, with an increase in all age groups from round 5 (26 January – 8 February 2021) (Supplementary Table S4). Vaccine hesitancy was uncommon with 1726 (0.8%, [0.7-0.8]) people reporting having refused or planning to refuse a vaccine when offered. Refusal was higher in those aged 25-34 (OR 1.25 [1.07-1.47]) compared to 35-44 years, and lower in the older age groups; it was lower in men ((0.83[0.75-0.91]) than women, and higher in those with lower income and educational level, and in Black, Mixed and Other ethnicities compared to white and Asian ethnicity. Vaccine hesitancy was also more likely in people who had a history of COVID-19, and in smokers. (Supplementary Table S5, Supplementary Figure S1) The most cited reasons for refusal (selected from a list) were concerns about the long-term health effects, side effects, or waiting to see whether it works, although the last reason was less frequently cited in round 6 than round 5 (Supplementary Figure S2).

### Antibody prevalence

The prevalence of antibodies (weighted to the adult population of England and adjusted for test characteristics) was 61.1% (95% CI 60.9-61.4), up from 6.6% (5.4-5.7) in round 4 (27 October – 10 November) and 13.9% (13.7-14.1) in round 5 (26 January – 8 February) (Table 1). There was a small variation in prevalence between regions with the lowest in the South West at 58.7% (57.9-59.5) and highest in London at 62.0% (61.4-62.7), but substantially more difference between LTLAs where prevalence ranged from 40.1% (35.6-44.9) to 78.7% (75.3-81.9) (Figure 2, Supplementary Table S2, Supplementary data file).

**Table 1 REACT-2:** Community prevalence of IgG antibodies to SARS-CoV-2 in adults in England, adjusted and weighted, June 2020 – May 2021.

|  | Total antibody positive | Total tests (with valid results) | Crude prevalence % (95% confidence intervals) | Adjusted and weighted<sup>1</sup> prevalence % (95% confidence intervals) |
| --- | --- | --- | --- | --- |
| Round 1 (20 Jun - 13 Jul 2020) | 5544 | 99908 | 5.6 (5.4-5.7) | 6.0 (5.8-6.1) |
| Round 2 (31 Jul – 13 Aug 2020) | 4995 | 105829 | 4.7 (4.6-4.9) | 4.8 (4.7-5.0) |
| Round 3 (15 - 28 Sep 2020) | 7037 | 159367 | 4.4 (4.3-4.5) | 4.4 (4.3-4.5) |
| Round 4 (27 Oct – 10 Nov 2020) | 8431 | 161537 | 5.2 (5.1-5.3) | 5.6 (5.4-5.7) |
| Round 5 (26 Jan – 8 Feb 2021) | 17059 | 154417 | 11.1 (10.9-11.2) | 13.9 (13.7-14.1) |
| Round 6 (12 – 25 May 2021) | 126981 | 207337 | 61.2 (61.0-61.5) | 61.1 (60.9-61.4) |

<sup>1</sup> Adjusted for test performance, and weighted to be representative of the adult population of England

Prevalence (adjusted and weighted) increased with age, from 35.8% (35.1-36.5) in those aged 18 – 24 years, to 95.3% (94.6-95.9) in people aged 75 years and over (Supplementary Table S2). Antibodies were 30% less likely to be detected in men than women (adjusted OR 0.69, 0.68-0.70), and were higher in people of Asian (OR 1.67 [1.58-1.77]), Black (1.55 [1.41-1.69]), mixed 1.17 [1.06-1.29] and other (1.37 [1.23-1.51]) ethnicities compared with white ethnicity (Supplementary Table S6, Supplementary Figure S3). There was a small gradient by deprivation; compared to the middle quintile of IMD, people living in the most deprived areas had lower (0.96, 0.93-1.00) and least deprived higher (1.06,1.04-1.09) odds of being antibody positive. Health workers, care workers, and other key workers, had higher odds of being antibody positive than other, non-key workers at 7.96 [7.49-8.47], 4.44 [3.89-5.07] and 1.29 [1.25-1.33] respectively. However, there were some public-facing workers with lower odds of having antibodies, including those working in hospitality (OR 0.69 [0.63-0.74]) and retail (0.71 [0.67-0.75]) compared to other workers (Supplementary Table S6).

### Antibody positivity following vaccination

The proportion of people testing positive following vaccination dose varied by number of doses, time since vaccination, age, sex and prior COVID-19 infection. Figure 3 shows the proportion of respondents who tested positive for antibodies (unadjusted and unweighted) by number of weeks since their first or second vaccine dose in people who had their second vaccination between 10 and 12 weeks after the first, or who had one vaccine less than 12 weeks earlier. It shows a rise in positivity after the first dose up to 4-5 weeks, and then a decline up to 11 weeks followed by a steep rise following a second dose. At each time point positivity was lower in men than women, in older than younger people, and higher in those with a history of COVID-19, and in those receiving Pfizer-BioNTech than AstraZeneca. (Figure 3 and Supplementary Figure S4)

**Figure 3:**
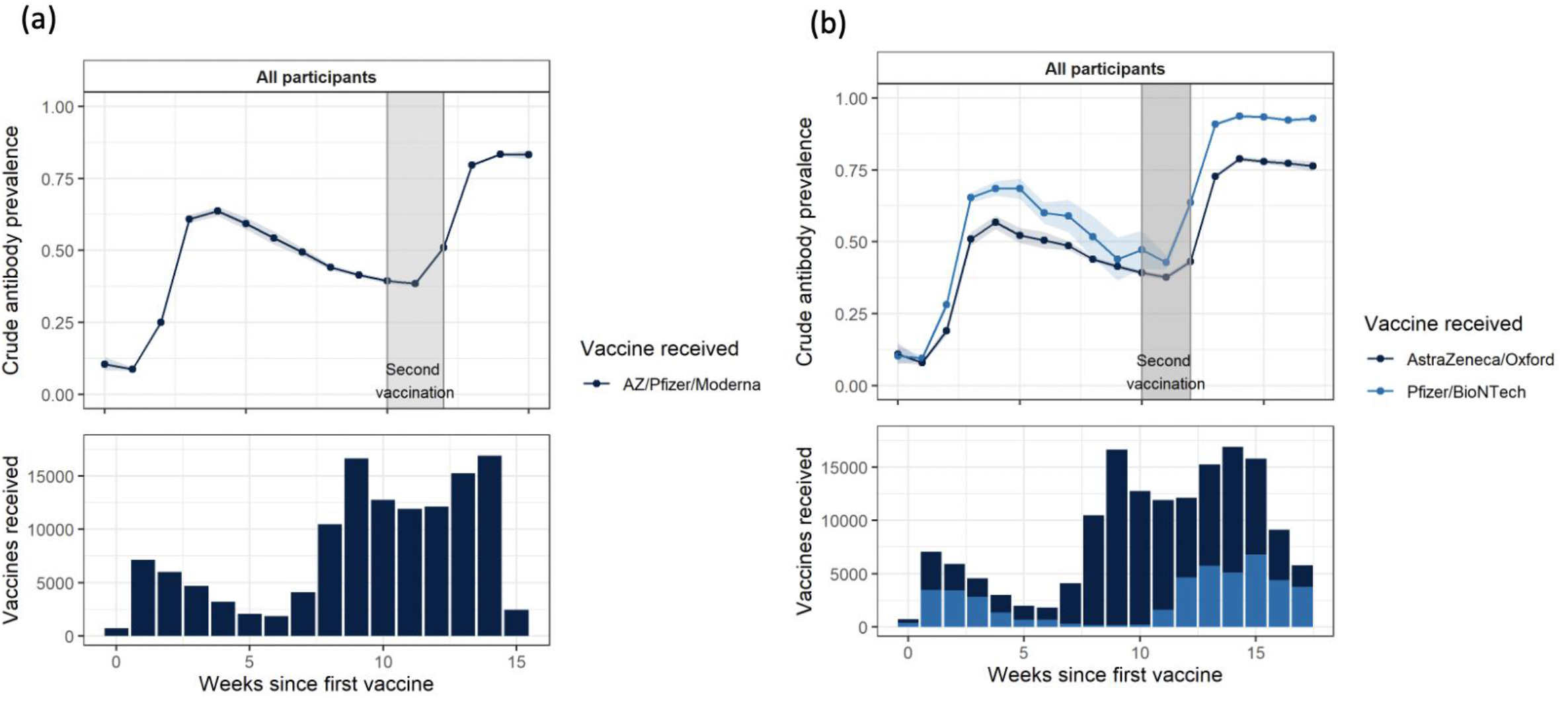
Percentage of respondents testing positive by weeks since dose (for people who had only one dose less than 12 weeks ago, or had second dose between 10 and 12 weeks after the first) (a) for all vaccine types, and (b) separately for Pfizer-BioNTech and AstraZeneca. Legend: The plots show the proportion of respondents who tested positive for antibodies according to time since their first and second vaccine dose. To ensure that we are comparing respondents who had their second vaccination at a similar time interval after the first, the plots show subsets of people who either 1) had their second vaccinations between 10 and 12 weeks after their first, or 2) have had one vaccine < 12 weeks ago. This captures 80% (168,877) of the 212,177 respondents who had received one or more vaccines. In (b) proportions are shown for AstraZeneca and Pfizer-BioNTech recipients; there were insufficient numbers to plot Moderna. Proportions are not adjusted for test performance.

Table 2 shows summary figures for antibody positivity 21 or more days following first, and 14 or more days following second doses separately for Pfizer-BioNTech, AstraZeneca and Moderna. After adjusting for test performance, the positivity after two doses was 100% (100-100) for Pfizer at all ages except 80 years and older when it was slightly lower at 97.8% (95.9-99.6) (Table 2a). For AstraZeneca adjusted positivity was over 90% up to age 69, and then 89.2% (88.5-89.9) in 70-79 year olds and 83.6% (78.5-88.3) in those aged 80 years and over (Table 2b). There was more variation following a single dose; after a single Pfizer-BioNTech dose, adjusted positivity ranged from 100.0% (91.1-100.0) in those aged 18-29 to 32.2% (18.2-51.1) in those aged 70 – 79 years (Table 2a). For AstraZeneca this was 72.2% (68.5-75.9) in the youngest and 46.2% (40.0-52.7) in the oldest age group (Table 2b). There were small numbers reporting Moderna vaccination with most being positive after a single dose (Table 2c).

**Table 2:** Antibody positivity (adjusted for test performance, not weighted) following vaccination by vaccine type, dose, time since dose and age.

| Age category | Positive | Total | Prevalence % | Lower CI | Upper CI |
| --- | --- | --- | --- | --- | --- |
| **One dose and >=21 days** |  |  |  |  |  |
| 18-29 | 99 | 117 | 100.0 | 91.1 | 100.0 |
| 30-39 | 127 | 177 | 84.8 | 76.3 | 92.1 |
| 40-49 | 196 | 335 | 68.8 | 62.4 | 75.0 |
| 50-59 | 362 | 842 | 50.1 | 46.1 | 54.2 |
| 60-69 | 385 | 995 | 44.9 | 41.3 | 48.6 |
| 70-79 | 11 | 39 | 32.3 | 18.2 | 51.1 |
| **Two doses and >=14 days** |  |  |  |  |  |
| 18-29 | 709 | 725 | 100.0 | 100.0 | 100.0 |
| 30-39 | 1198 | 1225 | 100.0 | 100.0 | 100.0 |
| 40-49 | 1622 | 1684 | 100.0 | 100.0 | 100.0 |
| 50-59 | 6493 | 6858 | 100.0 | 100.0 | 100.0 |
| 60-69 | 13461 | 14416 | 100.0 | 100.0 | 100.0 |
| 70-79 | 13273 | 14477 | 100.0 | 100.0 | 100.0 |
| 80+ | 1890 | 2288 | 97.8 | 95.9 | 99.6 |

(b) AstraZeneca
| Age category | Positive | Total | Prevalence % | Lower CI | Upper CI |
| --- | --- | --- | --- | --- | --- |
| **One dose and >=21 days** |  |  |  |  |  |
| 18-29 | 590 | 962 | 72.2 | 68.5 | 75.9 |
| 30-39 | 1055 | 1872 | 66.2 | 63.5 | 68.9 |
| 40-49 | 3089 | 6012 | 60.2 | 58.7 | 61.7 |
| 50-59 | 12482 | 29869 | 48.7 | 48.0 | 49.3 |
| 60-69 | 7590 | 20665 | 42.6 | 41.8 | 43.4 |
| 70-79 | 130 | 327 | 46.2 | 40.0 | 52.7 |
| **Two doses and >=14 days** |  |  |  |  |  |
| 18-29 | 310 | 346 | 100.0 | 100.0 | 100.0 |
| 30-39 | 481 | 535 | 100.0 | 100.0 | 100.0 |
| 40-49 | 703 | 838 | 99.4 | 96.2 | 100.0 |
| 50-59 | 3711 | 4680 | 93.8 | 92.4 | 95.2 |
| 60-69 | 13364 | 17170 | 92.1 | 91.3 | 92.8 |
| 70-79 | 16001 | 21209 | 89.2 | 88.5 | 89.9 |
| 80+ | 339 | 479 | 83.6 | 78.5 | 88.3 |

(c) Moderna
| Category | Positive | Total | Prevalence % | Lower CI | Upper CI |
| --- | --- | --- | --- | --- | --- |
| One dose and >=21 days |  |  |  |  |  |
| 18-29 | 6 | 6 | 100.0 | 81.4 | 100.0 |
| 40-49 | 261 | 286 | 100.0 | 100.0 | 100.0 |
| 50-59 | 9 | 11 | 96.9 | 61.3 | 100.0 |
| 60-69 | 5 | 7 | 84.4 | 41.6 | 100.0 |
| Two doses and >=14 days |  |  |  |  |  |
| 60-69 | 11 | 12 | 100.0 | 76.2 | 100.0 |

## Discussion

Our results confirm the rapid and widespread uptake of vaccination among adults across England and indicate a high antibody response among those who have had two doses of vaccine. Nonetheless, some groups still had relatively low vaccine coverage, in part explained by the ongoing prioritisation by age, with lowest vaccination rates at ages 18-24 years. However, lower coverage was also seen in men, people of Black, mixed or other ethnicity compared to Asian and white ethnicities, as well as those living in London, those in more deprived areas and those with lowest household income. Coverage was lower in occupational groups such as hospitality, retail, personal care, and other public facing roles, even after multiple adjustment including age and other factors, potentially placing these groups at increased risk of infection and ongoing transmission. Vaccination rates were also lower in those reporting suspected or confirmed prior COVID-19. In part this may be because in those with known infection it is recommended to wait four weeks before vaccination. However, it is also possible that those with prior infection may be less motivated to be vaccinated in the belief that they are protected.

Our overall estimate of antibody prevalence in mid-May 2021 was 61%, at a point when 69% of adults had received at least one dose of vaccine, with 39% having had two doses.(4) While lower than some other national estimates,(16,17) this is a large increase from the 14% we estimated at the end of January 2021 and shows the substantial impact of the vaccine programme. Specifically, our data show a clear increase in antibody response after second vaccination doses, providing strong support in favour of obtaining a second dose when offered. However, although viral neutralisation titres show a strong association with protection,(7)(18), the relationship between IgG positivity and risk of infection is still not fully understood and emerging variants, including delta, may require higher protective titres to achieve neutralisation.(19)

We confirmed in an independent population sample declines in antibody positivity following first vaccine dose, which we first reported in REACT 2 round 5 (26 January to 8 February 2021);(2) antibody positivity appeared to peak 3-4 weeks following first doses and then declined until after second doses were given. This may have implications for the duration between first and second doses particularly during a period of rapid epidemic growth.

Across regions there was a slightly lower prevalence in the South West where there were lower levels of prior infection, with the highest antibody prevalence found in regions with high rates of prior infection, London and the North West;(20) this is despite the lower vaccine coverage in London. Higher prior infection rates also resulted in higher antibody prevalence in non-white ethnic groups; however, there was a lower level of antibody prevalence in people living in deprived compared to more affluent areas, reflecting differential uptake of vaccines.

Our study has limitations. While we approached a random sample of the population, it is possible that differential non-response may have resulted in biased estimates of prevalence and vaccine uptake. Nonetheless, we achieved a large sample and relatively high participation (over 25%) amongst those invited to take part and weighted our findings to be representative of the population of England as a whole. Antibodies detected in this study reflected anti-S IgG in response to prior infection and vaccination, but this is only one part of the protective immune response, and therefore we cannot equate presence or absence of antibody with immunity. In addition, although offering distinct advantages in terms of costs, logistics and scale, the LFIA does have limitations. Due to limited sensitivity, it does not detect low levels of antibody which may result in lower antibody prevalence estimates than some other studies (21) and potentially be of clinical significance, for example, as a marker of protection against severe disease and/or hospitalisation; as people read their own test results there will be some classification errors including the missing of faint positive lines in people with low visual acuity. On the other hand, within-study comparisons, for example, by ethnicity or deprivation, should be relatively unaffected by test performance. Nonetheless we adjust our prevalence estimates for the performance of the test, based on previous validation work,(10) as well as presenting the observed proportions of test positives.

In summary our findings confirm the successful roll out of the vaccination programme in England with a concomitant high proportion of individuals having detectable antibodies. Those who have had two vaccines at the time of this study, including those over 65 years, had a very high proportion of antibody positivity which is likely to be associated with high levels of protection from severe disease, and possibly infection. Nonetheless, there remain some key groups with a lower prevalence of antibody, notably unvaccinated younger people, certain minority ethnic groups, those living in deprived areas and workers in some public-facing employment. Obtaining improved rates of vaccination in these groups is essential to achieving high levels of protection against the virus through population immunity.

## Data Availability

Data referred to in the manuscript are provided in tables in Supplementary material and through a linked data set.

https://github.com/mrc-ide/reactidd/tree/master/inst/extdata/react2_round6

## Contributors

HW, GC and PE designed the study and drafted the manuscript. MW and ST conducted the analyses. MW and ST verified the underlying data. HW, GC, CA, AD, CAD, PJD, SR, DA, WB, PE provided study oversight. AD and PE obtained funding. All authors have reviewed and approved the final manuscript

## Acknowledgements

HW is a National Institute for Health Research (NIHR) Senior Investigator and acknowledges support from NIHR Biomedical Research Centre, Imperial College NHS Trust, NIHR School of Public Health Research, NIHR Applied Research Collaborative North West London, and Wellcome Trust (UNS32973). GC is supported by an NIHR Professorship. WSB is the Action Medical Research Professor, AD is an NIHR senior investigator and DA and PE are Emeritus NIHR Senior Investigators. CAD and SR acknowledge support from MRC Centre for Global Infectious Disease Analysis (reference MR/R015600/1), jointly funded by the UK Medical Research Council (MRC) and the UK Foreign, Commonwealth & Development Office (FCDO), under the MRC/FCDO Concordat agreement. CAD acknowledges support from the NIHR HPRU in Emerging and Zoonotic Infectious (NIHR200907). SR acknowledges support from the NIHR Health Protection Research Unit (HPRU) in Respiratory Infections, Wellcome Trust (200861/Z/16/Z, 200187/Z/15/Z), and Centres for Disease Control and Prevention (US, U01CK0005-01-02). PE is Director of the MRC Centre for Environment and Health (MR/L01341X/1, MR/S019669/1). PE acknowledges support from the NIHR Imperial Biomedical Research Centre and the NIHR HPRUs in Chemical and Radiation Threats and Hazards and in Environmental Exposures and Health, the British Heart Foundation Centre for Research Excellence at Imperial College London (RE/18/4/34215), Health Data Research UK (HDR UK) and the UK Dementia Research Institute at Imperial (MC_PC_17114). We thank The Huo Family Foundation for their support of our work on COVID-19.

We thank key collaborators on this work -- Ipsos MORI: Stephen Finlay, John Kennedy, Kevin Pickering, Duncan Peskett, Sam Clemens and Kelly Beaver; Institute of Global Health Innovation at Imperial College London: Gianluca Fontana, Sutha Satkunarajah, Didi Thompson and Lenny Naar; the Imperial Patient Experience Research Centre and the REACT Public Advisory Panel; NHS Digital for access to the NHS Register.

## Data availability

Supporting data are available here.

## List of Tables and Figures

### Supplementary Tables

- Supplementary Table S1: Sample, registration and response rates, REACT-2 round 6, 12 – 25 May 2021
- Supplementary Table S2: Vaccination coverage (percent) and SARS-CoV-2 IgG antibody prevalence by socio-demographic characteristics, adjusted for test performance and weighted to the adult population of England
- Supplementary Table S3: Logistic regression models with vaccination status (one or more vaccine dose)
- Supplementary Table S4: Vaccine confidence and hesitancy by age, REACT-2 Round 5 (26 Jan – 8 Feb 2021) and Round 6 (12 – 25 May 2021)
- Supplementary Table S5: Vaccine hesitancy regression model
- Supplementary Table S6: Logistic regression for overall IgG antibody prevalence by key co-variates for all participants

### Supplementary Figures

- Supplementary Figure S1: Vaccine hesitancy: regression model
- Supplementary Figure S2: Reasons for actual or intended refusal of vaccine offer, REACT-2 Round 5 (26 Jan – 8 Feb 2021) and Round 6 (12 – 25 May 2021)
- Supplementary Figure S3: Logistic regression for overall IgG antibody prevalence by key co-variates
- Supplementary Figure S4: Percentage of respondents testing positive by weeks since dose by age, sex, prior COVID-19

## Footnotes

1 https://www.imperial.ac.uk/medicine/research-and-impact/groups/react-study/

