## Supplementary Tables and Figures for "Vaccine uptake and SARS-CoV-2 antibody prevalence among 207,337 adults during May 2021 in England: REACT-2 study"

### Supplementary Table S1: Sample, registration and response rates, REACT-2 round 6, 12 – 25 May 2021

| | Round 6<br>12 – 25 May 2021 | %<br>(of sampled) | %<br>(of registered) |
|---|---|---|---|
| Sample invited | 749212 | - | - |
| Registration - agreed to receive LFT test | 255750 | 34.0% | - |
| Symptom surveys done | 224244 | 29.9% | 87.7% |
| Attempted LFT test | 213620 | 28.5% | 83.5% |
| Valid (IgG positive or negative) test result | 207337 | 27.7% | 81.1% |

Supplementary Table S2 Vaccination coverage (percent) and SARS-CoV-2 IgG antibody prevalence by socio-demographic characteristics, adjusted for test performance and weighted to the adult population of England

| Category | Total unvaccinated | Total vaccinated | Percentage vaccinated | Antibody prevalence unvaccinated | Antibody prevalence vaccinated | Antibody prevalence all | Weighted percentage vaccinated | Weighted and adjusted antibody prevalence unvaccinated only | Weighted and adjusted antibody prevalence vaccinated only | Weighted and adjusted antibody prevalence all |
|---|---|---|---|---|---|---|---|---|---|---|
| **All participants** | | | | | | | | | | |
| All participants | 18552 | 187952 | 91.0 (90.9-91.1) | 13.6 (13.1-14.1) | 66.0 (65.8-66.2) | 61.2 (61.0-61.5) | 72.9 (72.7-73.0) | 14.9 (14.5-15.2) | 78.4 (78.1-78.7) | 61.1 (60.9-61.4) |
| **Sex** | | | | | | | | | | |
| Female | 11154 | 104976 | 90.4 (90.2-90.6) | 15.0 (14.3-15.7) | 69.1 (68.8-69.3) | 63.8 (63.6-64.1) | 75.3 (75.0-75.5) | 17.4 (16.8-17.9) | 83.0 (82.6-83.4) | 66.7 (66.4-67.1) |
| Male | 7396 | 82974 | 91.8 (91.6-92.0) | 11.6 (10.9-12.3) | 62.1 (61.7-62.4) | 57.9 (57.6-58.2) | 70.3 (70.0-70.6) | 12.7 (12.3-13.2) | 73.2 (72.8-73.6) | 55.2 (54.9-55.6) |
| **Age group** | | | | | | | | | | |
| 18-24 | 3451 | 1267 | 26.9 (25.6-28.1) | 14.1 (13.0-15.3) | 78.8 (76.5-81.0) | 31.5 (30.2-32.9) | 25.1 (24.5-25.6) | 17.0 (16.3-17.6) | 92.0 (90.7-93.3) | 35.8 (35.1-36.5) |
| 25-34 | 7342 | 3664 | 33.3 (32.4-34.2) | 11.3 (10.6-12.1) | 74.5 (73.1-75.9) | 32.4 (31.5-33.2) | 33.2 (32.8-33.7) | 12.0 (11.5-12.5) | 88.6 (87.6-89.5) | 37.5 (36.9-38) |
| 35-44 | 5064 | 9080 | 64.2 (63.4-65.0) | 11.9 (11.1-12.8) | 57.7 (56.7-58.7) | 41.3 (40.5-42.1) | 63.3 (62.8-63.8) | 14.5 (13.8-15.3) | 68.1 (67.3-68.9) | 48.4 (47.8-49) |
| 45-54 | 748 | 16772 | 95.7 (95.4-96.0) | 21.8 (19.0-24.9) | 57.9 (57.2-58.7) | 56.4 (55.7-57.1) | 95.1 (94.9-95.3) | 27.8 (25.4-30.3) | 68.6 (68.0-69.3) | 66.6 (66-67.2) |
| 55-64 | 1370 | 79167 | 98.3 (98.2-98.4) | 23.6 (21.5-26.0) | 56.5 (56.2-56.9) | 56.0 (55.6-56.3) | 98.0 (97.9-98.2) | 29.4 (25.4-33.8) | 67.0 (66.3-67.7) | 66.2 (65.6-66.9) |
| 65-74 | 534 | 70731 | 99.3 (99.2-99.3) | 20.2 (17.0-23.8) | 77.3 (77.0-77.7) | 76.9 (76.5-77.2) | 99.1 (99.0-99.2) | 24.8 (18.8-31.9) | 90.9 (90.3-91.5) | 90.2 (89.6-90.9) |
| 74+ | 43 | 7271 | 99.4 (99.2-99.6) | 23.3 (13.2-37.7) | 80.7 (79.8-81.6) | 80.3 (79.4-81.2) | 99.2 (99.1-99.3) | 41.0 (32.9-50.0) | 95.7 (95.1-96.3) | 95.3 (94.6-95.9) |
| **Ethnicity** | | | | | | | | | | |
| Asian | 948 | 5028 | 84.1 (83.2-85.0) | 18.7 (16.3-21.3) | 74.3 (73.1-75.5) | 65.3 (64.1-66.5) | 63.7 (62.9-64.5) | 21.1 (19.8-22.4) | 89.5 (88.4-90.5) | 64.5 (63.5-65.5) |
| Black | 378 | 1816 | 82.8 (81.1-84.3) | 31.5 (27.0-36.3) | 71.8 (69.7-73.8) | 64.9 (62.9-66.8) | 65.6 (64.3-66.8) | 35.7 (33.3-38.2) | 84.5 (82.8-86.2) | 67.8 (66.3-69.4) |
| Mixed | 463 | 1303 | 73.8 (71.7-75.8) | 15.3 (12.3-18.9) | 67.3 (64.7-69.8) | 53.7 (51.4-56.0) | 49.4 (47.4-51.4) | 20.7 (18.1-23.4) | 80.0 (76.8-83.2) | 50 (47.6-52.4) |
| Other | 278 | 1515 | 84.5 (82.7-86.1) | 23.7 (19.1-29.1) | 71.7 (69.4-73.9) | 64.0 (61.8-66.2) | 62.0 (59.9-64.0) | 23.8 (20.6-27.3) | 85.0 (82.0-87.9) | 61.5 (59-64.1) |

| | | | | | | | | | | |
|---|---|---|---|---|---|---|---|---|---|---|
| White | 16225 | 176959 | 91.6 (91.5-91.7) | 12.6 (12.1-13.1) | 65.6 (65.4-65.9) | 61.2 (60.9-61.4) | 74.3 (74.1-74.5) | 13.0 (12.6-13.3) | 77.4 (77.1-77.7) | 60.8 (60.6-61.1) |
| **Index of multiple deprivation (IMD) quintile** | | | | | | | | | | |
| 1 - most deprived | 2422 | 15916 | 86.8 (86.3-87.3) | 17.1 (15.6-18.6) | 64.8 (64.0-65.5) | 58.5 (57.7-59.2) | 67.4 (66.9-67.8) | 16.9 (16.2-17.7) | 77.8 (77.1-78.5) | 58 (57.4-58.6) |
| 2 | 3638 | 28200 | 88.6 (88.2-88.9) | 15.0 (13.9-16.2) | 65.3 (64.8-65.9) | 59.5 (59.0-60.1) | 69.4 (68.9-69.8) | 16.6 (15.8-17.3) | 78.2 (77.5-78.8) | 59.3 (58.7-59.9) |
| 3 | 4077 | 40173 | 90.8 (90.5-91.1) | 13.0 (12.0-14.0) | 65.7 (65.2-66.1) | 60.8 (60.3-61.2) | 73.4 (73.0-73.8) | 13.6 (12.9-14.4) | 78.6 (78.0-79.3) | 61.3 (60.7-61.8) |
| 4 | 4179 | 48441 | 92.1 (91.8-92.3) | 12.8 (11.9-13.9) | 66.4 (66.0-66.8) | 62.1 (61.7-62.5) | 75.8 (75.4-76.2) | 13.3 (12.6-14.1) | 78.8 (78.2-79.5) | 63 (62.4-63.5) |
| 5 - least deprived | 4236 | 55222 | 92.9 (92.7-93.1) | 11.9 (10.9-12.9) | 66.5 (66.1-66.9) | 62.6 (62.2-63.0) | 78.2 (77.8-78.6) | 12.8 (12.0-13.7) | 78.4 (77.7-79.0) | 64.1 (63.5-64.7) |
| **Region** | | | | | | | | | | |
| East Midlands | 2016 | 23959 | 92.2 (91.9-92.6) | 12.4 (11.0-13.9) | 64.8 (64.2-65.4) | 60.7 (60.2-61.3) | 75.3 (74.7-76.0) | 11.7 (10.6-12.8) | 76.6 (75.6-77.6) | 60.6 (59.7-61.5) |
| East of England | 2647 | 27118 | 91.1 (90.8-91.4) | 12.2 (11.0-13.5) | 65.2 (64.7-65.8) | 60.5 (59.9-61.0) | 74.6 (74.0-75.1) | 12.0 (11.1-13.0) | 77.5 (76.7-78.4) | 60.9 (60.1-61.7) |
| London | 2998 | 18281 | 85.9 (85.4-86.4) | 19.8 (18.5-21.3) | 72.8 (72.2-73.4) | 65.3 (64.6-65.9) | 64.5 (64.0-65.0) | 21.2 (20.4-22.1) | 84.5 (83.7-85.2) | 62 (61.4-62.7) |
| North East | 629 | 6978 | 91.7 (91.1-92.3) | 15.6 (13.0-18.6) | 64.1 (62.9-65.2) | 60.0 (58.9-61.1) | 73.2 (72.3-74.0) | 14.2 (12.7-15.8) | 76.0 (74.7-77.3) | 59.4 (58.2-60.5) |
| North West | 1959 | 22526 | 92.0 (91.7-92.3) | 16.7 (15.1-18.4) | 68.3 (67.7-68.9) | 64.1 (63.5-64.7) | 74.1 (73.5-74.6) | 18.5 (17.5-19.6) | 80.8 (80.0-81.6) | 64.6 (63.9-65.3) |
| South East | 3903 | 40870 | 91.3 (91.0-91.5) | 11.0 (10.1-12.0) | 64.5 (64.1-65.0) | 59.8 (59.4-60.3) | 74.4 (74.0-74.9) | 11.1 (10.3-11.9) | 76.5 (75.8-77.2) | 59.8 (59.1-60.4) |
| South West | 1675 | 17927 | 91.5 (91.1-91.8) | 8.4 (7.2-9.8) | 63.3 (62.6-64.0) | 58.5 (57.9-59.2) | 74.9 (74.3-75.5) | 9.0 (8.1-10.0) | 75.3 (74.4-76.2) | 58.7 (57.9-59.5) |
| West Midlands | 1651 | 17629 | 91.4 (91.0-91.8) | 14.2 (12.6-15.9) | 66.2 (65.5-66.9) | 61.7 (61.0-62.4) | 72.1 (71.5-72.7) | 16.8 (15.8-18.0) | 78.5 (77.6-79.4) | 61.3 (60.5-62.1) |
| Yorkshire and The Humber | 1074 | 12664 | 92.2 (91.7-92.6) | 12.1 (10.3-14.2) | 65.1 (64.2-65.9) | 60.9 (60.0-61.7) | 75.8 (75.2-76.3) | 12.8 (11.7-13.9) | 77.1 (76.2-78.0) | 61.5 (60.7-62.3) |

### Supplementary Table S3 Logistic regression models with vaccination status (one or more vaccine, yes or no) as the binary outcome variable

| Level | Unadjusted model | Adjusted on age and sex | Further adjusted on region, ethnicity, IMD, and healthcare or carehome worker status | Further adjusted on previous COVID-19 infection |
|---|---|---|---|---|
| **Sex** | | | | |
| Female [reference] | - | - | - | - |
| Male | 1.18 [1.15-1.22] | 0.83 [0.79-0.86] | 0.89 [0.85-0.94] | 0.89 [0.85-0.94] |
| **Age group** | | | | |
| 18-24 | 0.21 [0.20-0.23] | 0.21 [0.19-0.22] | 0.19 [0.17-0.20] | 0.18 [0.17-0.20] |
| 25-34 | 0.28 [0.27-0.29] | 0.28 [0.26-0.29] | 0.24 [0.23-0.26] | 0.24 [0.22-0.25] |
| 35-44 [reference] | - | - | - | - |
| 45-54 | 12.27 [11.36-13.26] | 12.35 [11.43-13.35] | 13.70 [12.54-14.95] | 13.67 [12.52-14.93] |
| 55-64 | 30.61 [28.82-32.50] | 31.01 [29.20-32.94] | 33.15 [30.79-35.70] | 32.53 [30.21-35.04] |
| 65-74 | 70.69 [64.85-77.06] | 72.05 [66.09-78.56] | 56.55 [47.63-67.14] | 54.19 [45.63-64.35] |
| 74+ | 97.63 [72.93-130.69] | 99.79 [74.54-133.60] | 207.26 [29.41-1460.58] | 197.45 [28.02-1391.40] |
| **Ethnicity** | | | | |
| White [reference] | - | - | - | - |
| Asian | 0.46 [0.43-0.49] | 1.10 [1.01-1.21] | 1.14 [1.02-1.28] | 1.13 [1.01-1.27] |
| Black | 0.42 [0.37-0.46] | 0.42 [0.36-0.49] | 0.41 [0.34-0.49] | 0.41 [0.34-0.49] |
| Mixed | 0.26 [0.23-0.29] | 0.77 [0.67-0.89] | 0.85 [0.71-1.01] | 0.86 [0.72-1.03] |
| Other | 0.46 [0.41-0.52] | 0.57 [0.48-0.68] | 0.76 [0.61-0.95] | 0.77 [0.62-0.97] |
| **Ethnicity (granular)** | | | | |
| English/Welsh/Scottish /Northern Irish/British [reference] | - | - | - | - |
| African | 0.30 [0.26-0.35] | 0.45 [0.38-0.55] | 0.37 [0.30-0.47] | 0.37 [0.30-0.47] |
| Arab | 0.24 [0.19-0.32] | 0.51 [0.35-0.75] | 0.48 [0.29-0.78] | 0.48 [0.29-0.78] |
| Bangladeshi | 0.19 [0.15-0.26] | 1.03 [0.73-1.45] | 1.18 [0.76-1.84] | 1.22 [0.78-1.90] |
| Caribbean | 0.49 [0.41-0.59] | 0.22 [0.17-0.28] | 0.22 [0.16-0.30] | 0.22 [0.16-0.30] |
| Chinese | 0.30 [0.25-0.36] | 0.68 [0.53-0.88] | 0.71 [0.52-0.97] | 0.69 [0.50-0.94] |
| Gypsy or Irish Traveller | 0.56 [0.22-1.44] | 0.44 [0.11-1.75] | 0.53 [0.11-2.52] | 0.53 [0.11-2.45] |
| Indian | 0.58 [0.53-0.64] | 1.26 [1.10-1.43] | 1.19 [1.02-1.39] | 1.17 [1.00-1.37] |
| Irish | 1.25 [1.06-1.48] | 0.85 [0.68-1.06] | 0.92 [0.71-1.19] | 0.93 [0.72-1.21] |
| Other Asian background | 0.37 [0.32-0.43] | 0.85 [0.70-1.04] | 0.77 [0.60-0.99] | 0.77 [0.59-0.99] |
| Other Black/African/Caribbean background | 0.48 [0.32-0.74] | 0.83 [0.47-1.45] | 1.15 [0.53-2.51] | 1.18 [0.53-2.60] |
| Other ethnic group | 0.46 [0.40-0.52] | 0.52 [0.43-0.63] | 0.68 [0.53-0.87] | 0.70 [0.54-0.90] |

| | | | | |
|---|---|---|---|---|
| Other Mixed/Multiple ethnic background | 0.24 [0.20-0.28] | 0.63 [0.49-0.80] | 0.72 [0.54-0.97] | 0.75 [0.56-1.00] |
| Other white background | 0.22 [0.21-0.23] | 0.37 [0.35-0.40] | 0.39 [0.36-0.43] | 0.40 [0.37-0.43] |
| Pakistani | 0.24 [0.20-0.28] | 0.72 [0.58-0.90] | 0.76 [0.56-1.01] | 0.78 [0.58-1.04] |
| Prefer not to say | 0.39 [0.34-0.44] | 0.27 [0.23-0.32] | | |
| White and Asian | 0.24 [0.20-0.29] | 0.82 [0.64-1.04] | 0.80 [0.60-1.07] | 0.80 [0.60-1.07] |
| White and Black African | 0.24 [0.17-0.34] | 0.47 [0.29-0.78] | 0.51 [0.28-0.93] | 0.52 [0.29-0.96] |
| White and Black Caribbean | 0.19 [0.15-0.24] | 0.71 [0.52-0.97] | 0.74 [0.52-1.07] | 0.78 [0.54-1.11] |
| **Region** | | | | |
| South East [reference] | - | - | - | - |
| East Midlands | 1.12 [1.06-1.19] | 1.19 [1.10-1.27] | 1.20 [1.10-1.31] | 1.19 [1.09-1.30] |
| East of England | 0.97 [0.92-1.02] | 1.00 [0.94-1.07] | 1.02 [0.95-1.11] | 1.02 [0.94-1.11] |
| London | 0.56 [0.53-0.59] | 0.72 [0.67-0.77] | 0.84 [0.77-0.91] | 0.86 [0.79-0.94] |
| North East | 1.04 [0.96-1.13] | 1.18 [1.05-1.32] | 1.11 [0.97-1.28] | 1.11 [0.97-1.28] |
| North West | 1.09 [1.03-1.15] | 1.14 [1.06-1.23] | 1.21 [1.11-1.33] | 1.23 [1.13-1.35] |
| South West | 1.03 [0.97-1.10] | 0.97 [0.89-1.04] | 0.93 [0.85-1.02] | 0.91 [0.83-1.00] |
| West Midlands | 1.01 [0.96-1.07] | 1.06 [0.98-1.14] | 1.10 [1.00-1.21] | 1.10 [1.00-1.21] |
| Yorkshire and The Humber | 1.13 [1.06-1.21] | 1.18 [1.08-1.29] | 1.23 [1.10-1.37] | 1.22 [1.09-1.36] |
| **Index of multiple deprivation (IMD) quintile** | | | | |
| 1 - most deprived | 0.65 [0.62-0.68] | 0.84 [0.78-0.90] | 0.82 [0.75-0.90] | 0.83 [0.76-0.91] |
| 2 | 0.78 [0.74-0.81] | 0.92 [0.86-0.97] | 0.94 [0.87-1.02] | 0.95 [0.88-1.03] |
| 3 [reference] | - | - | - | - |
| 4 | 1.18 [1.13-1.23] | 1.07 [1.01-1.13] | 1.05 [0.98-1.13] | 1.05 [0.98-1.12] |
| 5 - least deprived | 1.34 [1.29-1.40] | 1.20 [1.13-1.27] | 1.17 [1.09-1.26] | 1.17 [1.09-1.26] |
| **Gross household income** | | | | |
| 0-14,999 | 1.06 [0.99-1.13] | 0.62 [0.56-0.67] | 0.62 [0.54-0.71] | 0.62 [0.54-0.71] |
| 15,000-49,999 [reference] | - | - | - | - |
| 50,000-149,999 | 0.55 [0.53-0.57] | 1.11 [1.05-1.17] | 1.19 [1.12-1.27] | 1.19 [1.12-1.27] |
| >150,000 | 0.69 [0.63-0.75] | 1.19 [1.06-1.34] | 1.47 [1.28-1.68] | 1.48 [1.29-1.69] |
| **Previous case of COVID** | | | | |
| No COVID [reference] | - | - | - | - |
| COVID confirmed by test | 0.46 [0.43-0.49] | 0.68 [0.63-0.74] | 0.61 [0.55-0.67] | 0.61 [0.55-0.67] |
| COVID suspected by doctor | 0.46 [0.41-0.53] | 0.53 [0.45-0.63] | 0.58 [0.46-0.71] | 0.58 [0.46-0.71] |

| | | | | |
|---|---|---|---|---|
| COVID suspected by respondent | 0.36 [0.35-0.38] | 0.50 [0.48-0.53] | 0.57 [0.54-0.61] | 0.57 [0.54-0.61] |
| **BMI** | | | | |
| Normal weight [reference] | - | - | - | - |
| Obese | 1.78 [1.70-1.86] | 1.65 [1.55-1.75] | 1.63 [1.52-1.75] | 1.66 [1.55-1.78] |
| Overweight | 1.60 [1.54-1.66] | 1.14 [1.09-1.20] | 1.09 [1.03-1.16] | 1.11 [1.04-1.18] |
| Underweight | 0.53 [0.48-0.59] | 0.81 [0.69-0.96] | 0.75 [0.60-0.93] | 0.75 [0.60-0.93] |
| **Ever smoker** | | | | |
| Never cigarette smoker [reference] | - | - | - | - |
| Current cigarette smoker | 0.61 [0.58-0.64] | 0.78 [0.73-0.83] | 0.80 [0.73-0.86] | 0.80 [0.73-0.86] |
| Former cigarette smoker | 1.46 [1.41-1.51] | 0.95 [0.91-0.99] | 0.96 [0.91-1.01] | 0.97 [0.92-1.02] |
| Prefer not to say | 0.76 [0.70-0.83] | 0.63 [0.56-0.71] | 0.74 [0.63-0.86] | 0.75 [0.64-0.87] |
| **Keyworker/ work status** | | | | |
| Other worker [reference] | - | - | - | - |
| Care home worker | 2.88 [2.34-3.55] | 3.49 [2.71-4.50] | 3.93 [3.03-5.10] | 4.17 [3.20-5.43] |
| Healthcare worker | 4.26 [3.87-4.69] | 8.93 [8.01-9.95] | 9.42 [8.42-10.53] | 9.84 [8.79-11.02] |
| Other key worker | 1.02 [0.98-1.06] | 1.35 [1.28-1.42] | 1.34 [1.27-1.41] | 1.36 [1.29-1.44] |
| Other non worker | 4.01 [3.86-4.15] | 1.25 [1.18-1.32] | | |
| **Retail worker** | | | | |
| No [reference] | - | - | - | - |
| Yes | 0.67 [0.63-0.71] | 0.64 [0.58-0.70] | 0.77 [0.70-0.85] | 0.77 [0.70-0.85] |
| **Police, prisons, fire & rescue** | | | | |
| No [reference] | - | - | - | - |
| Yes | 1.32 [1.10-1.59] | 3.10 [2.50-3.86] | 3.59 [2.88-4.48] | 3.77 [3.02-4.72] |
| **Public transport worker** | | | | |
| No [reference] | - | - | - | - |
| Yes | 1.18 [1.00-1.38] | 0.80 [0.65-0.99] | 0.95 [0.76-1.18] | 0.97 [0.78-1.20] |
| **Education, school or nursey worker** | | | | |
| No [reference] | - | - | - | - |
| Yes | 1.12 [1.06-1.19] | 1.26 [1.17-1.36] | 1.59 [1.47-1.72] | 1.63 [1.50-1.76] |
| **Armed forces** | | | | |
| No [reference] | - | - | - | - |
| Yes | 1.45 [0.88-2.39] | 0.83 [0.42-1.65] | 0.86 [0.42-1.74] | 0.80 [0.39-1.64] |

| | | | | |
|---|---|---|---|---|
| **Other public facing role** | | | | |
| No [reference] | - | - | - | - |
| Yes | 0.94 [0.91-0.98] | 0.72 [0.68-0.76] | 0.82 [0.77-0.87] | 0.81 [0.77-0.86] |
| **Not currently required to work outside the home** | | | | |
| No [reference] | - | - | - | - |
| Yes | 0.80 [0.77-0.83] | 0.79 [0.75-0.83] | 1.02 [0.97-1.07] | 1.00 [0.95-1.05] |
| **Hospitality worker** | | | | |
| No [reference] | - | - | - | - |
| Yes | 0.41 [0.38-0.44] | 0.59 [0.52-0.67] | 0.72 [0.64-0.82] | 0.73 [0.64-0.82] |
| **Personal care, eg hairdresser, beauty therapist, personal trainer** | | | | |
| No [reference] | - | - | - | - |
| Yes | 0.43 [0.38-0.49] | 0.53 [0.44-0.64] | 0.61 [0.51-0.74] | 0.63 [0.52-0.76] |
| **Childcare worker** | | | | |
| No [reference] | - | - | - | - |
| Yes | 0.66 [0.55-0.80] | 1.14 [0.89-1.45] | 1.46 [1.14-1.87] | 1.44 [1.12-1.84] |

Supplementary Table S4: Vaccine confidence and hesitancy by age, REACT-2 Round 5 (26 Jan – 8 Feb 2021) and Round 6 (12 – 25 May 2021)

| | Round 5 | | | Round 6 | | |
|---|---|---|---|---|---|---|
| **Age_group** | **R5 Accepted / would accept vaccine** | **R5 Declined / would decline vaccine** | **R5 Don't know / prefer not to say** | **R6 Accepted / would accept vaccine** | **R6 Declined / would decline vaccine** | **R6 Don't know / prefer not to say** |
| **18-24** | 8,012 (83.7%, [82.9-84.4]) | 294 (3.1%, [2.7-3.4]) | 1,272 (13.3%, [12.6-14]) | 4,533 (91%, [90.2-91.8]) | 104 (2.1%, [1.7-2.5]) | 345 (6.9%, [6.3-7.7]) |
| **25-34** | 18,442 (83.3%, [82.8-83.8]) | 785 (3.5%, [3.3-3.8]) | 2,901 (13.1%, [12.7-13.6]) | 10,502 (89.1%, [88.5-89.6]) | 292 (2.5%, [2.2-2.8]) | 997 (8.5%, [8-9]) |
| **35-44** | 25,306 (87.4%, [87-87.8]) | 610 (2.1%, [1.9-2.3]) | 3,044 (10.5%, [10.2-10.9]) | 14,218 (92.2%, [91.8-92.6]) | 305 (2%, [1.8-2.2]) | 894 (5.8%, [5.4-6.2]) |
| **45-54** | 32,762 (92.4%, [92.2-92.7]) | 405 (1.1%, [1-1.3]) | 2,275 (6.4%, [6.2-6.7]) | 18,414 (97%, [96.8-97.3]) | 195 (1%, [0.9-1.2]) | 371 (2%, [1.8-2.2]) |
| **55-64** | 34,510 (95.8%, [95.5-96]) | 234 (0.6%, [0.6-0.7]) | 1,295 (3.6%, [3.4-3.8]) | 86,051 (98.5%, [98.4-98.5]) | 547 (0.6%, [0.6-0.7]) | 790 (0.9%, [0.8-1]) |
| **65-74** | 28,794 (98.1%, [97.9-98.2]) | 115 (0.4%, [0.3-0.5]) | 450 (1.5%, [1.4-1.7]) | 77,129 (99.3%, [99.3-99.4]) | 261 (0.3%, [0.3-0.4]) | 273 (0.4%, [0.3-0.4]) |
| **75+** | 10,489 (99%, [98.8-99.2]) | 33 (0.3%, [0.2-0.4]) | 71 (0.7%, [0.5-0.8]) | 7,987 (99.5%, [99.3-99.6]) | 22 (0.3%, [0.2-0.4]) | 18 (0.2%, [0.1-0.4]) |
| **All** | 158,315 (92%, [91.9-92.1]) | 2,476 (1.4%, [1.4-1.5]) | 11,308 (6.6%, [6.5-6.7]) | 218,834 (97.6%, [97.5-97.6]) | 1,726 (0.8%, [0.7-0.8]) | 3,688 (1.6%, [1.6-1.7]) |

Legend: Shows the number, percentage and 95% CI of respondents reporting actual or intended response to offer of a COVID-19 vaccine by age group and Round of REACT-2.

Supplementary Table S5 Vaccine hesitancy: regression model with binary outcome showing odds of responding I would / I did refuse the offer of a vaccine compared to all others (accept/ intend to accept, not sure, don't know)

| Covariate | Crude OR (95% CI) | OR adjusted for age (95% CI) | OR adjusted for age and sex (95% CI) |
|---|---|---|---|
| **Age group** | | | |
| 18-24 | 1.06 [0.84-1.32] | 1.06 [0.84-1.32] | 1.05 [0.84-1.31] |
| 25-34 | 1.26 [1.07-1.48] | 1.26 [1.07-1.48] | 1.25 [1.07-1.47] |
| 35-44 [reference] | NA [NA-NA] | NA [NA-NA] | NA [NA-NA] |
| 55-64 | 0.31 [0.27-0.36] | 0.31 [0.27-0.36] | 0.32 [0.27-0.36] |
| 65-74 | 0.17 [0.14-0.2] | 0.17 [0.14-0.2] | 0.17 [0.14-0.2] |
| 75+ | 0.14 [0.09-0.21] | 0.14 [0.09-0.21] | 0.14 [0.09-0.21] |
| **Sex** | | | |
| Female [reference] | NA [NA-NA] | NA [NA-NA] | NA [NA-NA] |
| Male | 0.74 [0.67-0.82] | 0.83 [0.75-0.91] | 0.83 [0.75-0.91] |
| **Level of education** | | | |
| No qualification | 0.95 [0.79-1.15] | 1.38 [1.14-1.67] | 1.39 [1.14-1.68] |
| GCSE [reference] | NA [NA-NA] | NA [NA-NA] | NA [NA-NA] |
| Post-GCSE qualification | 1.21 [1.06-1.39] | 0.97 [0.84-1.12] | 0.98 [0.85-1.13] |
| Degree or above | 1.14 [1-1.3] | 0.83 [0.73-0.95] | 0.84 [0.73-0.96] |
| Other | 1.18 [0.97-1.44] | 1.46 [1.2-1.78] | 1.49 [1.22-1.81] |
| **Ethnicity** | | | |
| White [reference] | NA [NA-NA] | NA [NA-NA] | NA [NA-NA] |
| Asian | 1.16 [0.89-1.51] | 0.80 [0.62-1.05] | 0.81 [0.62-1.06] |
| Black | 2.59 [1.93-3.47] | 2.09 [1.56-2.81] | 2.11 [1.57-2.83] |
| Mixed | 2.67 [1.92-3.71] | 1.59 [1.14-2.21] | 1.58 [1.14-2.21] |
| Other | 2.31 [1.64-3.23] | 1.91 [1.36-2.68] | 1.93 [1.37-2.7] |
| **Region** | | | |
| South East [reference] | NA [NA-NA] | NA [NA-NA] | NA [NA-NA] |
| East Midlands | 0.94 [0.78-1.13] | 0.95 [0.79-1.14] | 0.95 [0.79-1.14] |

| | | | |
|---|---|---|---|
| East of England | 1.14 [0.97-1.34] | 1.13 [0.96-1.33] | 1.13 [0.96-1.33] |
| London | 1.51 [1.28-1.78] | 1.29 [1.09-1.52] | 1.29 [1.09-1.53] |
| North East | 1.12 [0.86-1.47] | 1.12 [0.85-1.46] | 1.12 [0.85-1.46] |
| North West | 1.24 [1.04-1.47] | 1.25 [1.05-1.48] | 1.25 [1.05-1.48] |
| South West | 1.03 [0.85-1.25] | 1.07 [0.88-1.29] | 1.07 [0.88-1.29] |
| West Midlands | 1.14 [0.94-1.37] | 1.14 [0.94-1.37] | 1.14 [0.94-1.38] |
| Yorkshire and The Humber | 1.12 [0.9-1.38] | 1.14 [0.92-1.41] | 1.13 [0.92-1.41] |
| **Past COVID-19** | | | |
| No COVID-19 [reference] | NA [NA-NA] | NA [NA-NA] | NA [NA-NA] |
| COVID-19 confirmed by test | 3.11 [2.63-3.68] | 2.46 [2.07-2.91] | 2.44 [2.06-2.89] |
| COVID-19 suspected by doctor | 3.76 [2.76-5.14] | 3.1 [2.27-4.24] | 3.06 [2.24-4.19] |
| COVID-19 suspected by respondent | 4.06 [3.63-4.54] | 3.15 [2.8-3.53] | 3.15 [2.81-3.54] |
| **Gross household income** | | | |
| 0-14,999 | 1.67 [1.43-1.96] | 1.94 [1.65-2.27] | 1.91 [1.63-2.24] |
| 15,000-49,999 [reference] | NA [NA-NA] | NA [NA-NA] | NA [NA-NA] |
| 50,000-149,999 | 0.85 [0.73-0.98] | 0.59 [0.51-0.68] | 0.59 [0.51-0.69] |
| >150,000 | 0.5 [0.32-0.78] | 0.35 [0.23-0.55] | 0.36 [0.23-0.56] |
| **IMD (deprivation) Quintile** | | | |
| 1 - most deprived | 2.05 [1.76-2.38] | 1.79 [1.54-2.08] | 1.79 [1.54-2.08] |
| 2 - | 1.31 [1.13-1.52] | 1.21 [1.05-1.41] | 1.21 [1.05-1.4] |
| 3 [reference] | NA [NA-NA] | NA [NA-NA] | NA [NA-NA] |
| 4 - | 0.84 [0.73-0.97] | 0.87 [0.76-1.01] | 0.87 [0.76-1.01] |
| 5 - least deprived | 0.59 [0.51-0.69] | 0.63 [0.54-0.73] | 0.63 [0.54-0.73] |
| **Current smoker** | | | |
| Not current cigarette smoker [reference] | NA [NA-NA] | NA [NA-NA] | NA [NA-NA] |
| Current cigarette smoker | 2.00 [1.74-2.31] | 1.62 [1.41-1.87] | 1.63 [1.42-1.88] |
| Prefer not to say | 2.13 [1.54-2.93] | 1.77 [1.28-2.44] | 1.78 [1.29-2.45] |

Supplementary Table S6 Logistic regression for overall IgG antibody prevalence by key co-variates , all participants

| Level | Unadjusted model | Adjusted on age and sex | Further adjusted on region, ethnicity and IMD |
|---|---|---|---|
| **Sex** | | | |
| Female [reference] | - | - | - |
| Male | 0.78 [0.77-0.79] | 0.70 [0.68-0.71] | 0.69 [0.68-0.71] |
| **Age group** | | | |
| 35-44 [reference] | - | - | - |
| 18-24 | 0.65 [0.61-0.70] | 0.64 [0.60-0.69] | 0.66 [0.61-0.70] |
| 25-34 | 0.68 [0.65-0.72] | 0.67 [0.64-0.71] | 0.67 [0.64-0.71] |
| 45-54 | 1.84 [1.76-1.92] | 1.86 [1.78-1.95] | 1.91 [1.83-2.00] |
| 55-64 | 1.81 [1.74-1.87] | 1.85 [1.79-1.92] | 1.92 [1.85-1.99] |
| 65-74 | 4.72 [4.55-4.90] | 4.93 [4.74-5.12] | 5.16 [4.96-5.36] |
| 74+ | 5.81 [5.44-6.21] | 6.11 [5.71-6.53] | 6.44 [6.02-6.89] |
| **Ethnicity** | | | |
| White [reference] | - | - | - |
| Asian | 1.19 [1.13-1.26] | 1.67 [1.58-1.77] | 1.54 [1.46-1.64] |
| Black | 1.17 [1.07-1.28] | 1.55 [1.41-1.69] | 1.41 [1.29-1.55] |
| Mixed | 0.74 [0.67-0.81] | 1.17 [1.06-1.29] | 1.12 [1.01-1.24] |
| Other | 1.13 [1.03-1.24] | 1.37 [1.23-1.51] | 1.24 [1.12-1.37] |
| **Region** | | | |
| South East [reference] | - | - | - |
| East Midlands | 1.04 [1.01-1.07] | 1.03 [1.00-1.07] | 1.05 [1.01-1.08] |
| East of England | 1.03 [1.00-1.06] | 1.03 [1.00-1.07] | 1.04 [1.01-1.07] |
| London | 1.26 [1.22-1.31] | 1.47 [1.42-1.52] | 1.42 [1.37-1.47] |
| North East | 1.01 [0.96-1.06] | 1.02 [0.97-1.07] | 1.05 [1.00-1.11] |
| North West | 1.20 [1.16-1.24] | 1.21 [1.17-1.25] | 1.24 [1.20-1.29] |
| South West | 0.95 [0.92-0.98] | 0.92 [0.89-0.95] | 0.94 [0.90-0.97] |
| West Midlands | 1.08 [1.04-1.12] | 1.09 [1.05-1.13] | 1.10 [1.06-1.14] |
| Yorkshire and The Humber | 1.04 [1.00-1.08] | 1.03 [0.99-1.08] | 1.05 [1.01-1.10] |
| **Index of multiple deprivation (IMD) quintile** | | | |
| 1 - most deprived | 0.91 [0.88-0.94] | 1.01 [0.97-1.05] | 0.96 [0.93-1.00] |
| 2 | 0.95 [0.92-0.98] | 1.01 [0.98-1.04] | 0.97 [0.94-1.01] |
| 3 [reference] | - | - | - |
| 4 | 1.06 [1.03-1.08] | 1.03 [1.00-1.06] | 1.04 [1.01-1.06] |
| 5 - least deprived | 1.08 [1.05-1.11] | 1.04 [1.01-1.07] | 1.06 [1.04-1.09] |
| **Previous case of COVID** | | | |

| | | | |
|---|---|---|---|
| COVID confirmed by test | 6.80 [6.31-7.32] | 10.43 [9.66-11.26] | 10.18 [9.43-11.00] |
| COVID suspected by doctor | 1.62 [1.46-1.79] | 2.11 [1.90-2.34] | 2.09 [1.88-2.32] |
| COVID suspected by respondent | 1.24 [1.20-1.28] | 1.71 [1.66-1.77] | 1.70 [1.64-1.76] |
| No COVID [reference] | - | - | - |
| **BMI** | | | |
| Normal weight [reference] | - | - | - |
| Obese | 0.96 [0.94-0.98] | 1.00 [0.98-1.03] | 1.03 [1.00-1.06] |
| Overweight | 1.00 [0.98-1.02] | 0.99 [0.97-1.02] | 1.01 [0.98-1.03] |
| Underweight | 0.86 [0.79-0.94] | 0.89 [0.81-0.98] | 0.87 [0.79-0.96] |
| **Ever smoker** | | | |
| Never cigarette smoker [reference] | - | - | - |
| Current cigarette smoker | 0.65 [0.63-0.67] | 0.74 [0.72-0.77] | 0.76 [0.73-0.79] |
| Former cigarette smoker | 1.08 [1.06-1.10] | 0.99 [0.97-1.01] | 1.01 [0.99-1.03] |
| Prefer not to say | 0.89 [0.84-0.95] | 0.89 [0.84-0.95] | 0.90 [0.85-0.96] |
| **Gross household income** | | | |
| 15,000-49,999 [reference] | - | - | - |
| >150,000 | 0.77 [0.73-0.82] | 1.13 [1.06-1.20] | 1.09 [1.02-1.16] |
| 0-14,999 | 0.87 [0.84-0.91] | 0.76 [0.73-0.79] | 0.75 [0.73-0.78] |
| 50,000-149,999 | 0.78 [0.76-0.80] | 1.08 [1.05-1.11] | 1.08 [1.05-1.11] |
| **Ethnicity (granular)** | | | |
| English/Welsh/Scottish/Northern Irish/British [reference] | - | - | - |
| African | 1.19 [1.06-1.34] | 1.77 [1.57-2.01] | 1.62 [1.43-1.84] |
| Arab | 0.83 [0.66-1.06] | 1.34 [1.04-1.72] | 1.21 [0.94-1.55] |
| Bangladeshi | 0.97 [0.74-1.26] | 1.97 [1.50-2.59] | 1.73 [1.31-2.27] |
| Caribbean | 1.19 [1.04-1.38] | 1.34 [1.16-1.55] | 1.21 [1.04-1.40] |
| Chinese | 0.83 [0.71-0.97] | 1.16 [0.99-1.37] | 1.08 [0.92-1.27] |
| Gypsy or Irish Traveller | 1.74 [0.81-3.73] | 1.92 [0.86-4.29] | 1.97 [0.88-4.41] |
| Indian | 1.32 [1.23-1.42] | 1.78 [1.65-1.92] | 1.63 [1.51-1.77] |
| Irish | 1.21 [1.11-1.32] | 1.17 [1.07-1.28] | 1.10 [1.00-1.20] |
| Other Asian background | 1.13 [1.00-1.27] | 1.56 [1.37-1.77] | 1.43 [1.26-1.63] |
| Other Black/African/Caribbean background | 0.82 [0.60-1.11] | 1.20 [0.87-1.65] | 1.10 [0.80-1.52] |
| Other ethnic group | 1.18 [1.06-1.31] | 1.37 [1.23-1.53] | 1.24 [1.11-1.39] |
| Other Mixed/Multiple ethnic background | 0.71 [0.61-0.84] | 1.09 [0.92-1.29] | 1.03 [0.87-1.22] |
| Other white background | 0.71 [0.68-0.75] | 0.99 [0.95-1.04] | 0.93 [0.89-0.98] |
| Pakistani | 1.13 [0.97-1.32] | 1.89 [1.61-2.22] | 1.76 [1.50-2.07] |

| | | | |
|---|---|---|---|
| Prefer not to say | 0.82 [0.74-0.91] | 0.91 [0.82-1.01] | |
| White and Asian | 0.76 [0.65-0.89] | 1.23 [1.04-1.45] | 1.19 [1.01-1.40] |
| White and Black African | 0.94 [0.69-1.28] | 1.44 [1.05-1.99] | 1.34 [0.97-1.84] |
| White and Black Caribbean | 0.62 [0.51-0.77] | 1.09 [0.88-1.35] | 1.05 [0.84-1.30] |
| **Keyworker/ work status** | | | |
| Other worker [reference] | - | - | - |
| Care home worker | 4.43 [3.90-5.04] | 4.26 [3.73-4.85] | 4.44 [3.89-5.07] |
| Healthcare worker | 7.25 [6.84-7.69] | 7.85 [7.39-8.34] | 7.96 [7.49-8.47] |
| Other key worker | 1.15 [1.11-1.18] | 1.26 [1.22-1.29] | 1.29 [1.25-1.33] |
| Other non worker | 2.21 [2.16-2.25] | 1.13 [1.11-1.16] | 1.14 [1.11-1.17] |
| **Retail worker** | | | |
| No [reference] | - | - | - |
| Yes | 0.75 [0.71-0.79] | 0.71 [0.68-0.75] | 0.71 [0.67-0.75] |
| **Police, prisons, fire & rescue** | | | |
| No [reference] | - | - | - |
| Yes | 1.10 [0.97-1.24] | 1.41 [1.24-1.60] | 1.45 [1.28-1.64] |
| **Public transport worker** | | | |
| No [reference] | - | - | - |
| Yes | 0.84 [0.75-0.93] | 0.91 [0.81-1.02] | 0.85 [0.76-0.95] |
| **Education, school or nursery worker** | | | |
| No [reference] | - | - | - |
| Yes | 1.02 [0.98-1.06] | 0.95 [0.91-0.99] | 0.96 [0.92-1.00] |
| **Armed forces** | | | |
| No [reference] | - | - | - |
| Yes | 0.70 [0.52-0.95] | 0.77 [0.56-1.05] | 0.80 [0.58-1.10] |
| **Other public facing role** | | | |
| No [reference] | - | - | - |
| Yes | 0.75 [0.73-0.77] | 0.78 [0.76-0.81] | 0.80 [0.78-0.82] |
| **Not currently required to work outside the home** | | | |
| No [reference] | - | - | - |
| Yes | 0.73 [0.71-0.75] | 0.73 [0.71-0.75] | 0.71 [0.69-0.73] |
| **Hospitality worker** | | | |
| No [reference] | - | - | - |
| Yes | 0.63 [0.58-0.68] | 0.68 [0.63-0.74] | 0.69 [0.63-0.74] |
| **Personal care, eg hairdresser, beauty therapist, personal trainer** | | | |

| | | | |
|---|---|---|---|
| No [reference] | - | - | - |
| Yes | 0.73 [0.65-0.82] | 0.72 [0.64-0.82] | 0.74 [0.65-0.83] |
| **Childcare worker** | | | |
| No [reference] | - | - | - |
| Yes | 1.13 [0.97-1.31] | 1.13 [0.97-1.31] | 1.14 [0.98-1.34] |

Supplementary Figure S1: Vaccine hesitancy: regression model

Plot with binary outcome showing odds of responding I would / I did refuse the offer of a vaccine compared to all others (accept/ intend to accept, not sure, don't know); data in table S5

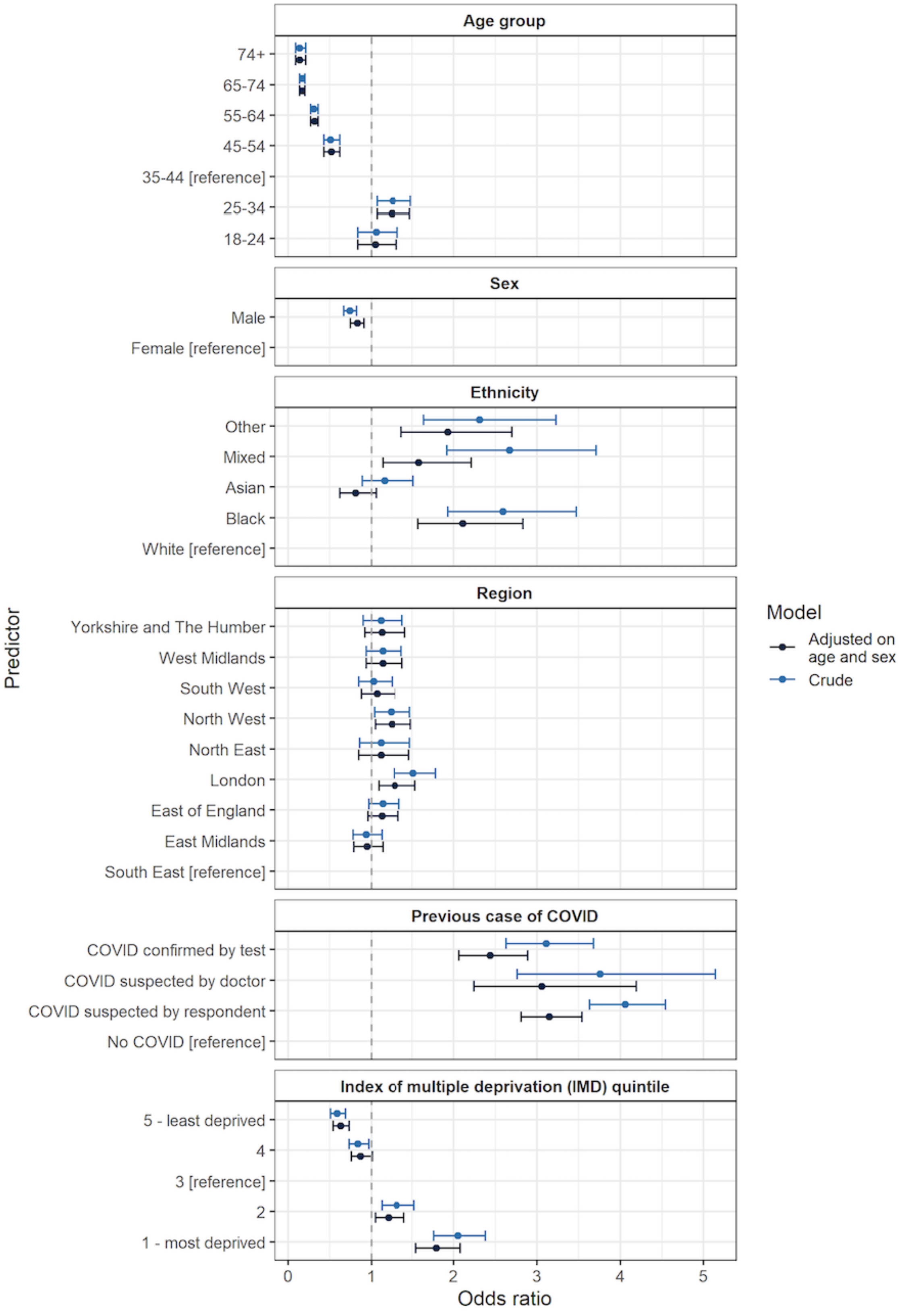

Supplementary Figure S2: Reasons for actual or intended refusal of vaccine offer, REACT-2 Round 5 (26 Jan – 8 Feb 2021) and Round 6 (12 – 25 May 2021)

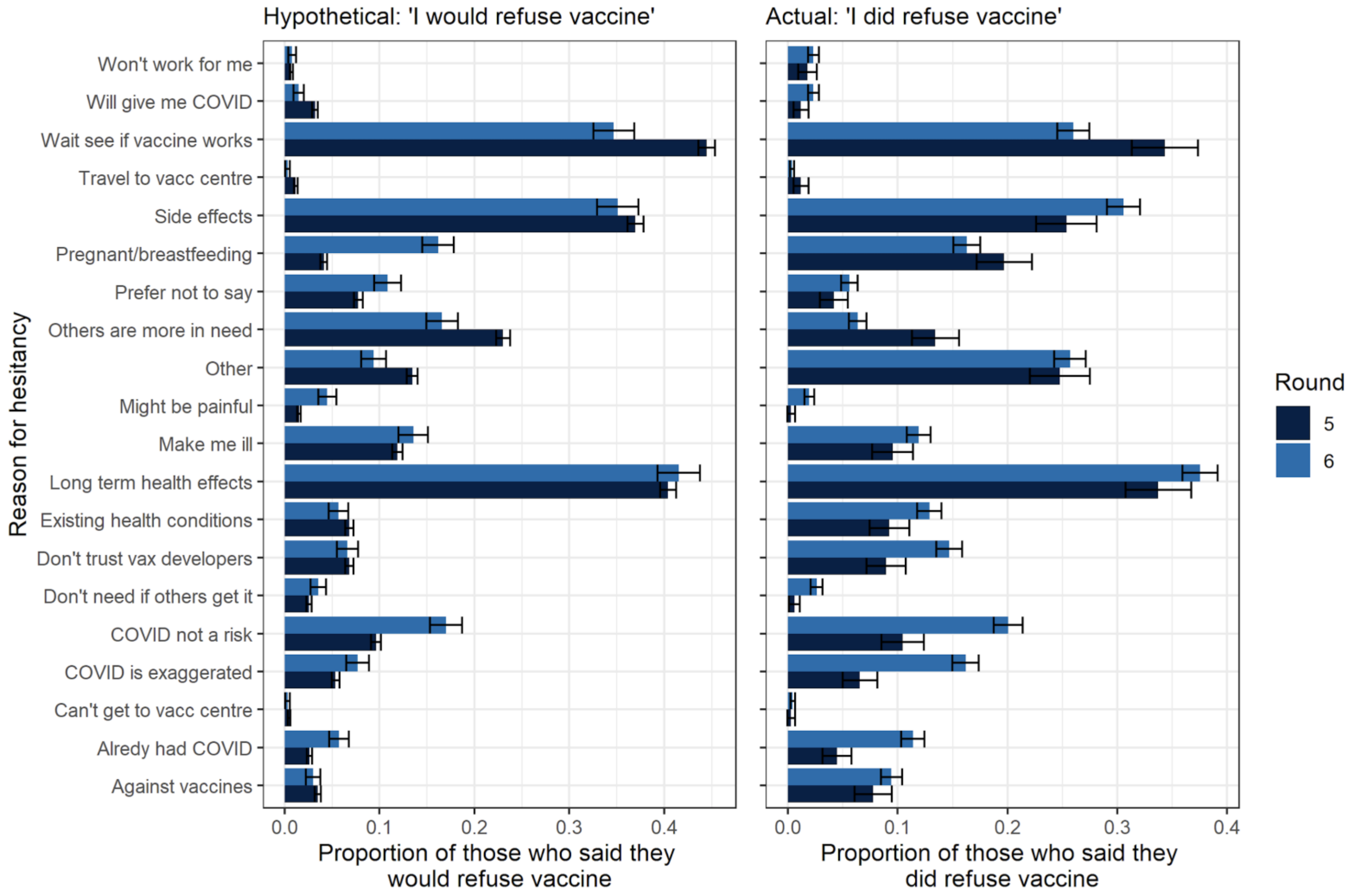

### Supplementary Figure S3 Logistic regression for overall IgG antibody prevalence by key covariates

The plot shows odds ratios for logistic regression models with antibody status as the binary outcome variable (one or more doses received, y/n) and a range of covariates as explanatory variables. Data for this chart are in Supplementary Table S6

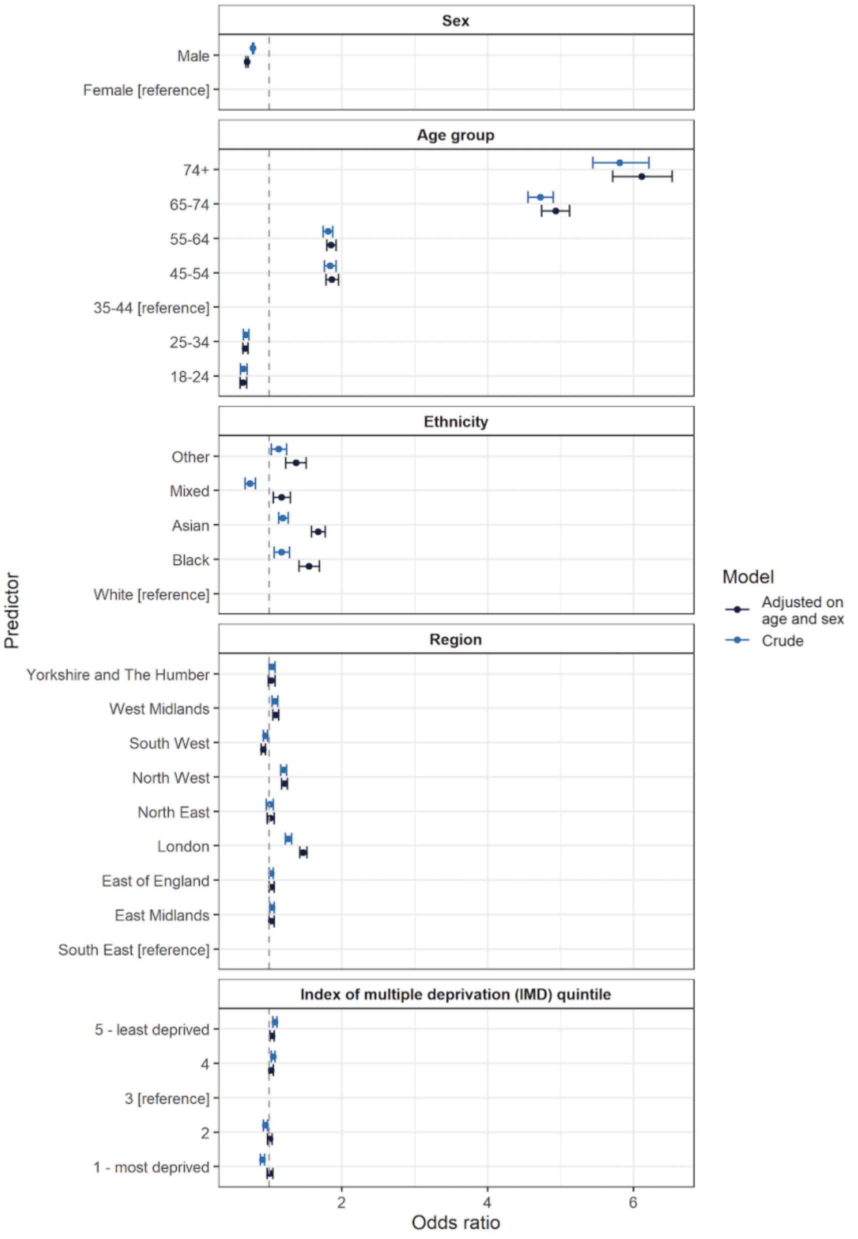

### Supplementary Figure S4 Percentage of respondents testing positive by weeks since dose by age, sex, prior COVID-19

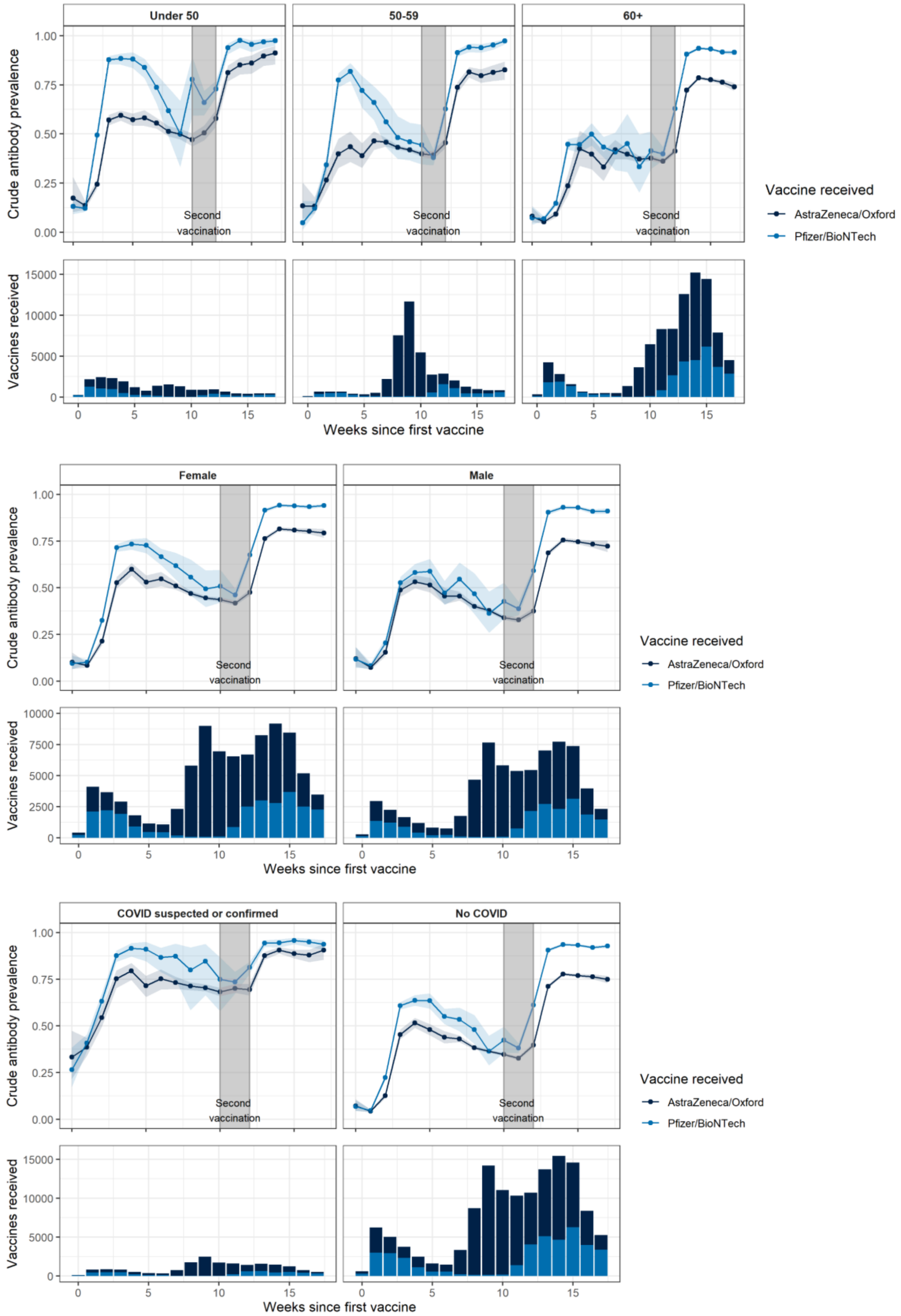

Legend: The plots show the proportion of respondents who tested positive for antibodies according to time since from their first and second vaccine dose. To ensure that we are comparing respondents who had their second vaccination at a similar time interval after the first, the plots show subsets of people who either 1) had their second vaccinations between 10 and 12 weeks after their first, or 2) have had one vaccine < 12 weeks ago. This captures 80% (168,877) of the 212,177 respondents who had received one or more vaccines. Proportions are shown for AstraZeneca and Pfizer recipients. Prior COVID-19 includes those with suspected or confirmed COVID-19 infection. Proportions are not adjusted for test performance
